# A versatile, fast and unbiased method for estimation of gene-by-environment interaction effects on biobank-scale datasets

**DOI:** 10.1101/2021.04.24.21255884

**Authors:** Mohammad Khan, Matteo Di Scipio, Conor Judge, Nicolas Perrot, Michael Chong, Shihong Mao, Shuang Di, Walter Nelson, Jeremy Petch, Guillaume Paré

## Abstract

Current methods to evaluate gene-by-environment (GxE) interactions on biobank-scale datasets are limited. MonsterLM enables multiple linear regression on genome-wide datasets, does not rely on parameters specification and provides unbiased estimates of variance explained by GxE interaction effects. We applied MonsterLM to the UK Biobank for eight blood biomarkers (N=325,991), identifying significant genome-wide interaction variance with waist-to-hip ratio for five biomarkers, with variance explained by interactions ranging from 0.11 to 0.58. 48% to 94% of GxE interaction variance can be attributed to variants without significant marginal association with the phenotype of interest. Conversely, for most traits, >40% of interaction variance was explained by less than 5% of genetic variants. We observed significant improvements in polygenic score prediction with incorporation of GxE interactions in four biomarkers. Our results imply an important contribution of GxE interaction effects, driven largely by a restricted set of variants distinct from loci with strong marginal effects.

## Introduction

Identifying gene-by-environment (GxE) interactions is difficult because individual interaction effects are expected to be small^1^, the multiple hypothesis burden is considerable^2,3^, and the sample sizes needed are correspondingly large^4^. Most previous analyses have focused on identifying interactions with variants marginally associated with a phenotype of interest^5,6^. Hitherto, methods developed to estimate the overall effect of these interactions rely on variance component methods, due to the predictor (*m*) > observation (*n*) problem, where single nucleotide polymorphisms (SNPs) (*m*) vastly outnumber the participants (*n*)^7,8^. These methods are advantageous for smaller datasets; however, they can be limited when applied to larger datasets due to computational burden^7^. Furthermore, variance component methods depend on strong assumptions about the underlying genetic model and often require *a priori* specification of parameters and/or hyper-parameters, such as polygenicity, minor allele frequency (MAF), and linkage disequilibrium (LD) dependence^9–13^. While never formally tested in the context of GxE interactions, it has previously been shown these assumptions can lead to important biases in heritability estimates ^9–11,14–18^. Novel methods are thus needed to enable fast and unbiased calculations of the variance explained (R^2^) by GxE interactions in large samples, on multiple traits and without the need for genetic model assumptions.

Our method is similar to the generalized random effects (GRE) model^19^, building on the observation that the multiple regression coefficient of determination can be used to accurately estimate heritability^19^. Extending this observation to include an environmental exposure variable and computing the interactions between genotypes and the environmental exposure allows us to examine the variance explained by genetic interactions with an environmental exposure. Using linear regression across the genome presents a key problem: there are far more SNPs (*m*) than participants (*n*) in genome-wide studies, and thus it becomes difficult to estimate heritability and interaction variance^20,21^. By partitioning the genome into non-overlapping regions, it becomes possible to estimate genome-wide interactions with environmental exposures by reducing *m* within each region to a size where *m* < *n*. However, partitioning the genome into large blocks still presents challenges. First, LD spillage at the junction of blocks can theoretically inflate heritability estimates if many such junctions exist^9^. Second, any residual population stratification effects would be amplified if heritability at each region is overestimated and this effect is expected to be proportional to the number of blocks^22^. Third, computing prediction R^2^ on large blocks with high dimensionality can be slow. By using the conjugate gradient method^23^ with graphics processing unit (GPU) acceleration^24^, it is possible to perform multiple linear regression modelling efficiently on large (25,000 SNPs) blocks (Supplementary Table 1). The potential for residual population stratification effects and LD spills are minimized as only approximately 60 blocks are typically needed for genome-wide analyses and variants are LD-pruned. A block size of 25,000 SNPs also ensures that *n* > 10*m* for accurate estimations.

**Table 1.** Comparison of current methods estimating GxE contributions to MonsterLM

| **Method** | **Description** | **Advantages** | **Disadvantages** | **MonsterLM** |
| --- | --- | --- | --- | --- |
| StructLMM<sup>26</sup> | Evaluates interaction variance for multiple environmental factors with a single SNP. | Fast, robust model for a variety of different environmental exposures. | Limited to interaction effects of only a single SNP or genotype. | Analyzes variance explained by interactions genome-wide (after LD-pruning). |
| CGI-GREML<sup>27</sup> | Uses a mix of parametrized models and restricted likelihood methods to estimate variance explained by GxE. | Well-structured for identifying GxE interactions with categorical exposures. | >250 Likelihood Ratio Tests; Slow; Cannot use continuous traits. | Can analyze continuous traits without categorizing them; Quick, efficient Wald-test/CI for R2 of interaction effects. |
| GxEMM<sup>28</sup> | Linear mixed model method to detect GxE interactions across the genome and a single exposure. | Multiple parametrizations available to efficiently model GxE interaction effects. | Small sample size only; Minimal number of SNPs. | Can analyze a large sample size with many SNPs through partition of genotype matrix & Conjugate Gradient method. |
| GRSxE<sup>29</sup> | Method to detect total GxE interactions with a Gene-Risk Score. | Estimates the GxE contribution for all possible environmental factors with SNPs. | Assumes each SNP interacts equally with E. | Accounts for the unique interaction effect of each SNP with E. |
| LEMMA<sup>30</sup> | Linear mixed model method to detect GxE interactions across the genome and an estimated linear combination of exposures. | Considers the impact of overlapping environmental exposures when computing total GxE contributions across the genome. | Requires parametrization and is dependent on model specification; Uses an estimated linear combination of exposures, assuming all E's interact with the same SNPs. | Tests for specific interaction with E rather than a linear combination of E. |

We propose a novel method, MonsterLM, to estimate the proportion of variance explained by GxE interactions for continuous traits, in a fast, accurate, efficient and unbiased manner on biobank-scale datasets (*N*>300,000). We hypothesized that GxE interactions contribute significantly to complex trait variance. Our objective was to quantify and characterize these contributions for continuous traits. We illustrate an overview of our computational analyses in Figure 1.

**Figure 1.**
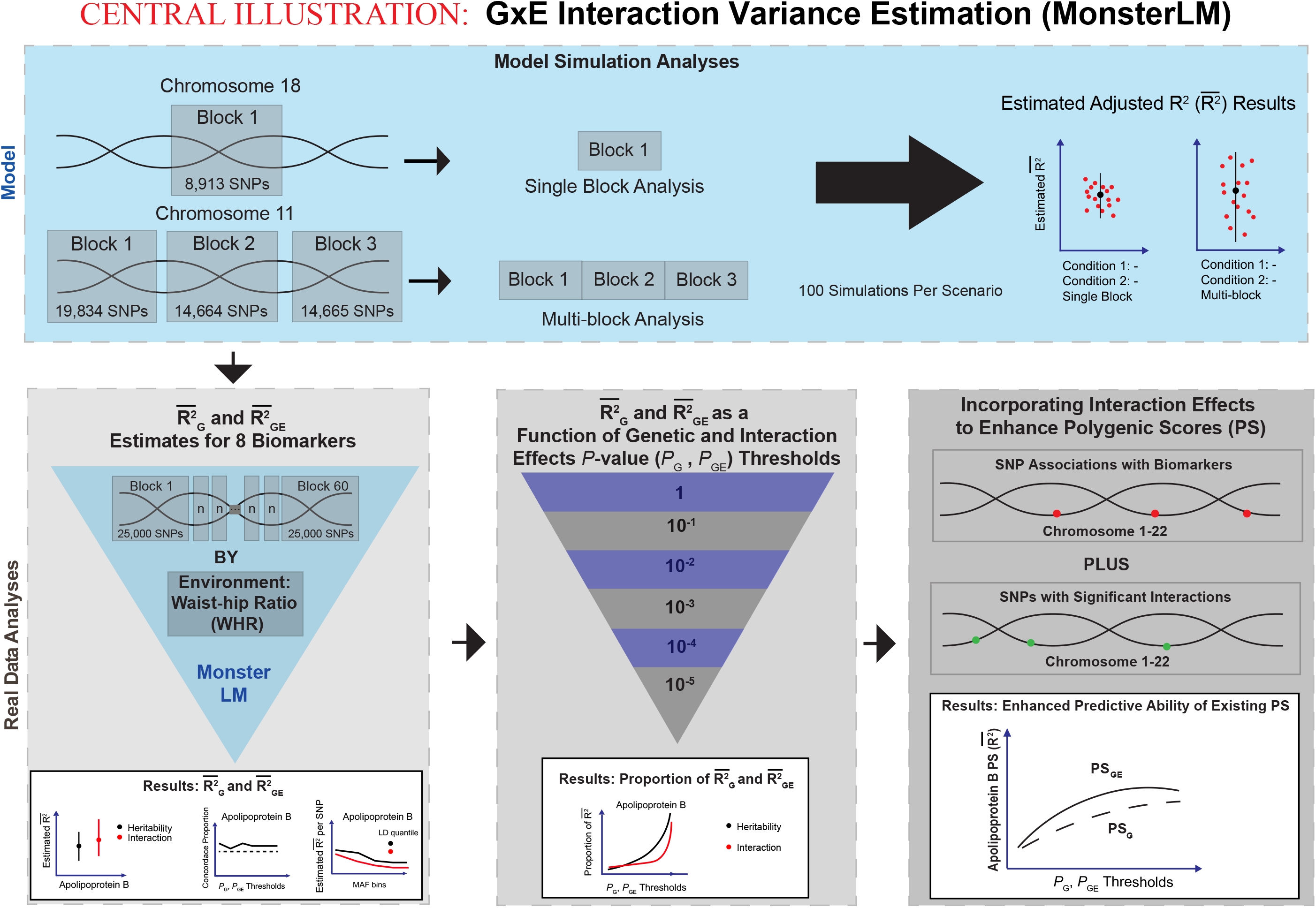
Summary of Gene-by-Environment (GxE) analysis conducted with MonsterLM. Initial simulation studies were conducted to verify the properties of MonsterLM; simulated phenotypes with known values for variance explained were regressed under varying SNP partitioning and interaction structure conditions to ensure robust estimations **(blue panel)**. Real trait analyses were conducted with UK Biobank data **(grey panels)**. Genome-wide SNP heritability estimates with and without waist-hip-ratio (WHR) interactions revealed significant interaction effects for five of eight biomarkers and were further assessed with a directionality of effects and stratification analysis **(bottom left panel)**. The model was further explored by recovering genotype and interaction variance explained through partitioning SNPs based on genotype and interaction univariate regressions thus providing insights into the model’s architecture **(bottom middle panel)**. Lastly, sequential incorporation of subsets of SNPs with significant *P*_*GE*_ derived from univariate interaction regressions of the genotype SNPs on their respective traits revealed modest improvements of polygenic scores (*PS*_*G*_,*PS*_*GE*_) in four of the five biomarkers tested **(bottom right panel)**.

## Methods

### UK Biobank

The UK Biobank is a large population-based study which includes over 500,000 participants living in the United Kingdom ^15,32^. Men and women aged 40–69 years were recruited between 2006 and 2010, and extensive phenotypic and genotypic data was collected. We selected 325,991 unrelated British individuals from the UK Biobank with both genotype and biomarker data for inclusion in the analysis. This study used genetic variants from the ‘V3’ release of the UK Biobank data including those present in the Haplotype Reference Consortium and 1000 Genomes panels with imputation quality greater than 0.7, no deviation from Hardy-Weinberg equilibrium (*P*>1×10^−10^) and minor allele frequency greater than 1% ^15^. Genotype data were filtered by removing highly correlated SNPs with a LD r^2^ value of more than 0.9 and removing SNPs with a MAF of less than 0.01, as the focus of this report is on common variants. After quality control filtering, there remained 1,031,135 SNPs and 325,991 individuals. Raw genotypes were normalized to have a mean of zero and variance of one. For the current analysis we examined eight biomarkers including Apolipoprotein B, Bilirubin, Total Cholesterol, C-reactive protein (CRP), HbA1c, HDL-Cholesterol, LDL-Cholesterol, Triglycerides and the environmental exposure, waist-to-hip ratio (WHR).

For secondary analyses, we randomly partitioned the UK Biobank participants into two sets: a discovery set containing 80% of the participants used for model building and a validation set containing the remaining 20% of the participants. This was done to remove the potential for overfitting that can occur when using derived models on the same datasets for prediction purposes^33^.

### MonsterLM Estimations of Variance Explained by GxE Effects

The standard linear model for a phenotypic trait (*Y*) when an interaction term is included can be expressed as:

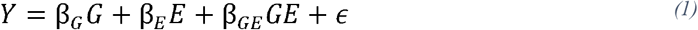

Where *G* is the genotype matrix, *E* is the environmental exposure, *GE* is the Hadamard product between each genotype and environmental exposure, resulting in a matrix with the same dimensionality as *G*. The betas (β) represent the true marginal effects associated with their respective term. To account for covariate effects such as age, sex, *E*, and population stratification we first regress *Y* onto the covariates and the first twenty genetic principal components and extract the residuals of the model (*y*_*residuals*_). The residuals (*y*_*residuals*_) become our phenotype used for analyses in MonsterLM:

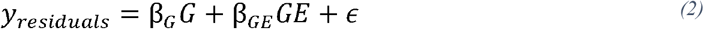

Both phenotype and environmental exposures are quantile normalized after residualization, such that mean is zero and variance one. Through residualization of the environmental exposure, we can leave *E* out of the model. For simplicity, we denote the augmented matrix of *G* and *GE* as *U* with dimension *n*×2*m*, where *n* is number of participants included and *m* is number of SNPs:

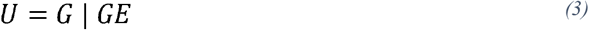

And *y*_*residuals*_ becomes:

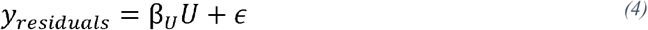

The MonsterLM method enables multiple linear regression on biobank-scale datasets by parallelizing the calculation of least squares regression, including the interaction terms, between the genotypes and environmental factors. The calculation is done such that the only practical limitation is the inversion of the *U* matrix, but without any restriction on *n*. This limitation is circumvented using the conjugate gradient method and GPU acceleration^24^. Importantly, MonsterLM requires neither parametrization nor assumptions regarding the genetic architecture of traits analyzed (such as polygenicity of effects, MAF and LD dependence). Genotypic data was partitioned into blocks with a maximal size of 25,000 SNPs (*m*) to minimize LD spillage between blocks and to optimize speed of the matrix calculation.

Given a quantitative trait *Y*, the least squares estimate for 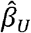, the estimated effects vector, corresponding to the genotype and GxE interaction is:

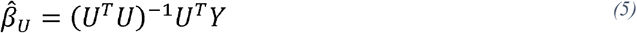

After computing 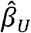 using conjugate gradient, the predicted values of Y denoted as *ŷ*, can be computed as:

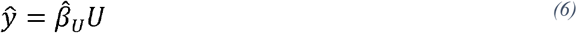

This same method can be applied if we use the genotype matrix only (*G*) instead of *U* to compute 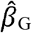 and *ŷ*. Once *ŷ* is calculated for each block (with and without interactions), we calculate the variance explained for the full model (*U*) and the model without interactions. Since r^2^ is a biased estimator, the adjusted r^2^ 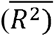 is used as our estimate for variance explained. Then, to calculate the interaction variance explained 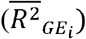 we compute the difference in 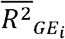 as:

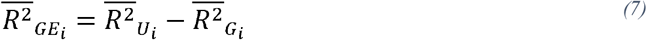

Since we remove SNPs in very high LD (r^2^ > 0.9), the remaining variant set will not be highly correlated. We can then estimate the total contribution of variance by genome-wide environment interaction 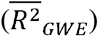 by taking the sum over all blocks:

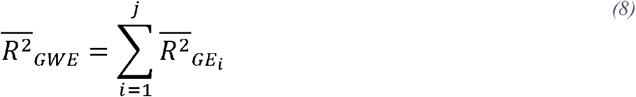

Where *j* is the number of partitioned blocks used for analysis (i.e. 60 blocks for current analyses) and *i* is the index of the current block.

The 95% confidence (CI) of the 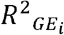 term can be estimated for each block using asymptotic properties described by Graf and Alf ^34^. The asymptotic variance for the difference between the 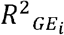 of two models is given by:

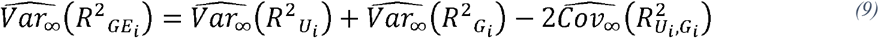

Where:

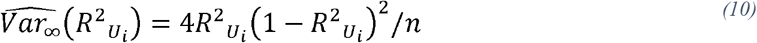

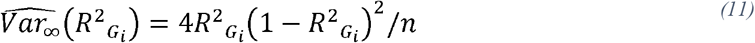

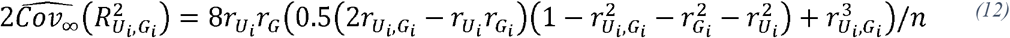

And:

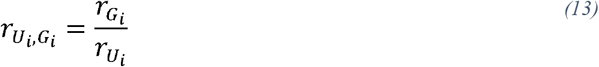

The 95% CI for a single block can then be derived using the Wald estimate:

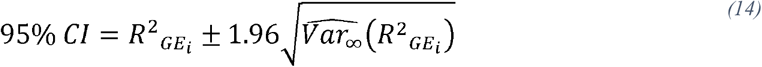

To estimate the 95% CI for our 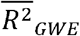 estimate, we calculate the total asymptotic variance as the sum of the individual variances 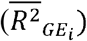 for each block, but since our estimates use 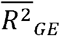 to estimate GxE interactions, we also adjust the variance of 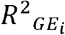 similarly to obtain the adjusted asymptotic variance of 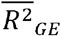, which is then used to calculate the 95% CIs.

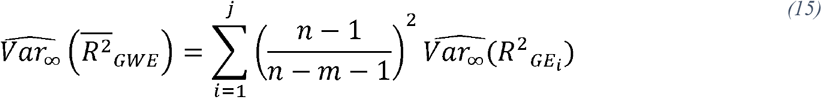

Where *n* is the number of samples and *m* is the number of SNPs tested per block *i*. With the total asymptotic variance estimated, we calculate the 95% CI for the 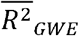 as:

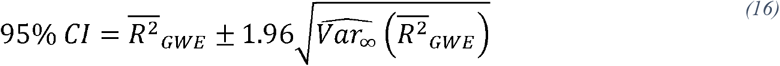

### Simulations to Validate the MonsterLM Method

We tested MonsterLM with simulations using UK Biobank genotypes filtered as described above. We used chromosome 18 to generate a single block of 8,913 SNPs (smallest block allowing for efficient simulations). We then simulated the true, unobserved effects (β_*G*_, β_*E*_, β_*GE*_) from a normal distribution, assuming 20% of SNPs have a marginal effect associated with the simulated trait of interest, *Y*_*sim*_ (i.e. β_*G*_ ≠ 0). We further assumed that 10% of the causal SNPs (i.e. 2% of total SNPs) have an interaction effect (i.e. β_*GE*_ ≠ 0). The values were chosen based on similar estimates with heritability of WHR through MonsterLM. The error was sampled from an independent and identically distributed normal distribution. The simulated trait (*Y*_*sim*_)was then computed as:

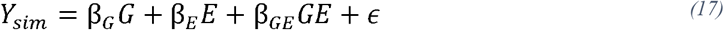

We divided the above case into three scenarios. The first scenario considered that *E* was not dependent on *G* and the genetic and interaction effects for all SNPs were randomly generated from a standard normal distribution. The next two scenarios considered that *E* was dependent on *G*. In these scenarios, *E* was simulated to have 20% of its variance explained by *G* (i.e. heritability), as WHR was observed to have similar heritability empirically. Scenario 2 further assumed that the genetic effects could be zero when the interaction effect was non-zero for a specific SNP *i* (β_*G,i*_ = 0, β_*GE,i*_ ≠ 0) and the SNPs explaining *E* were the same as the SNPs with an interaction effect. Scenario 3 assumed that both the genetic and interaction effects were non-zero for a specific SNP *i* (β_*G*,i_ ≠ 0, β_*GE*,i_ ≠ 0) and that the SNPs explaining *E* were not the same as the SNPs with an interaction effect. To ensure realistic scenarios were simulated, we varied the variance of the normal distributions to achieve pre-specified genetic, environment and interaction effects. The heritability 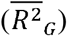 was set to 0.025, variance explained by the environmental exposure 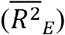 was set to 0.2 and variance explained by the interactions 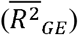 was set to 0.005. We also considered 3 multi-block scenarios identical to the above scenarios; whereby chromosome 11 was split into 3 blocks of roughly 15,000 SNPs each. Each block had 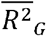 set to 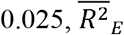 to 0.2, and 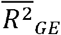 set to 0.005, such that the variance explained by interactions across the whole chromosome 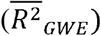 was 0.015.

### Directionality of Effects Analysis

After computing 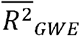 for our eight biomarkers, we tested whether direction of effect was concordant between marginal and interaction regression coefficients for each SNP. Concordant direction of effects is defined as when 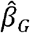 has the same sign (+/+, -/-) as 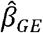 for a single SNP and its associated interaction. Discordant direction of effects is defined as when the 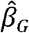 and 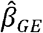 have a different sign (+/-, -/+) for a single SNP and its associated interaction. We used a subset of 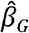 and 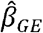 coefficients that were in low LD (r^2^ < 0.1) and computed the direction of effect concordance for this subset. We then plotted the sign concordance twice: first as a function of 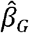 *P* - values (*P*_*G*_), then as a function of 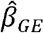 *P* - values (*P*_*GE*_), which were computed from association of single SNPs and their respective interaction on the biomarker traits. Two-proportion *Z*-tests were used to compare the proportion of directionally concordant marginal and interaction effects for each biomarker in each threshold compared to a null count at a proportion of 0.50.

### Stratification of Estimates by MAF and LD

SNPs were stratified by MAF and LD score into a total of 20 bins: 5 MAF bins (0.01≤0.1, 0.1<MAF≤0.2, 0.2<MAF≤0.3, 0.3<MAF≤0.4, and 0.4<MAF≤0.5) and 4 LD score quantiles (0<LD≤0.25, 0.25<LD≤0.50, 0.50<LD≤0.75, and 0.75<LD≤0.9). MAF and LD score were calculated using a subset of 5000 participants from the UKBiobank. We then computed the variance explained 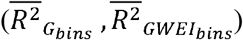 and divided each estimate by the total number of SNPs in each bin to get an 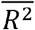 per SNP value that was compared between bins and to the total genetic and interaction variance estimates 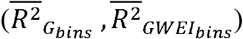.

### Polygenic Scores Analysis

To calculate polygenic scores (*PS*) without interactions (*PS*_*G*_), we first selected SNPs based on univariate *P*_*G*_ derived from regression of each variant with biomarker concentration from the discovery set. We then combined the selected SNPs into a single block from the discovery set, and applied MonsterLM regression to obtain the multiple linear regression coefficients 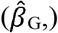. Using these coefficients, we calculated the *PS*_*G*_ in the validation set as:

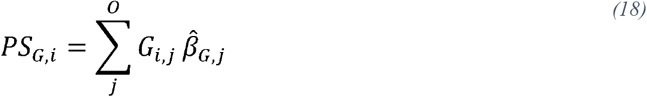

Where *PS*_*G,i*_ is the individual polygenic score of participant *i, j* is the SNP number and *O* represents the total number of SNPs included in this analysis. We then evaluated the predictiveness of each *PS*_*G*_ using 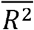 in the validation set. We repeated the same process for four univariate *P*_*G*_ thresholds (10^−2^, 10^−3^, 10^−4^, 10^−5^) for each biomarker.

We define *PS*_*GE*_ as the *PS* with GxE interactions included. To include GxE interactions, we selected significant interactions based on *P*_*GE*_ obtained from regressing each variant and its associated GxE interaction with biomarker concentration in the discovery set. These interactions are selected from the subset of SNPs included in polygenic scores without interactions. The interactions passing the univariate *P*_*GE*_ thresholds (10^−2^, 10^−3^, 10^−4^, 10^−5^) were then included with the SNPs to create a single block. We applied MonsterLM regression to obtain the multiple linear regression coefficients 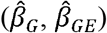. Using these coefficients, we calculate the *PS*_*GE*_ as:

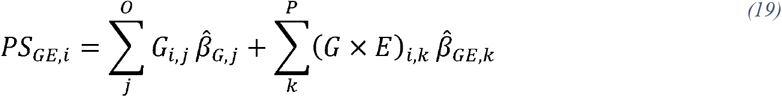

Where *PS*_*GE*_,_*i*_ is the polygenic score with interactions incorporated for participant *i*, summed over each SNP (*j*) and, if included, its associated interaction (*k*). *O* represents the SNPs included in the *PS*_*GE*_, while *P* represents the interactions included, a subset of *O*. As with the *PS*_*G*_, we evaluated the predictiveness of each polygenic score using 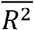 in the validation set. We repeated for all pairwise combinations of the four *P*_*G*_ thresholds and the four *P*_*GE*_ thresholds, resulting in 16 *PS*_*GE*_ for each biomarker.

## Results

### Simulation results

We conducted 100 simulations for each of the three scenarios (Figure 2A). On average, the 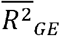 was observed to be close to 0.005, the true underlying 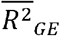 that was predefined for interactions. We compared the estimated 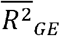 to the true 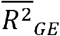 and found that the difference in 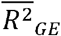 was not significant (*P*>0.05). After verifying MonsterLM for a single block, we conducted 100 simulations using three contiguous blocks from chromosome 11 under the same three scenarios. The 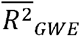 was observed to be close to 0.015, the true 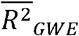 set for interactions (Figure 2B). Our calculated 95% CIs were also well-calibrated for our simulated data.

**Figure 2.**
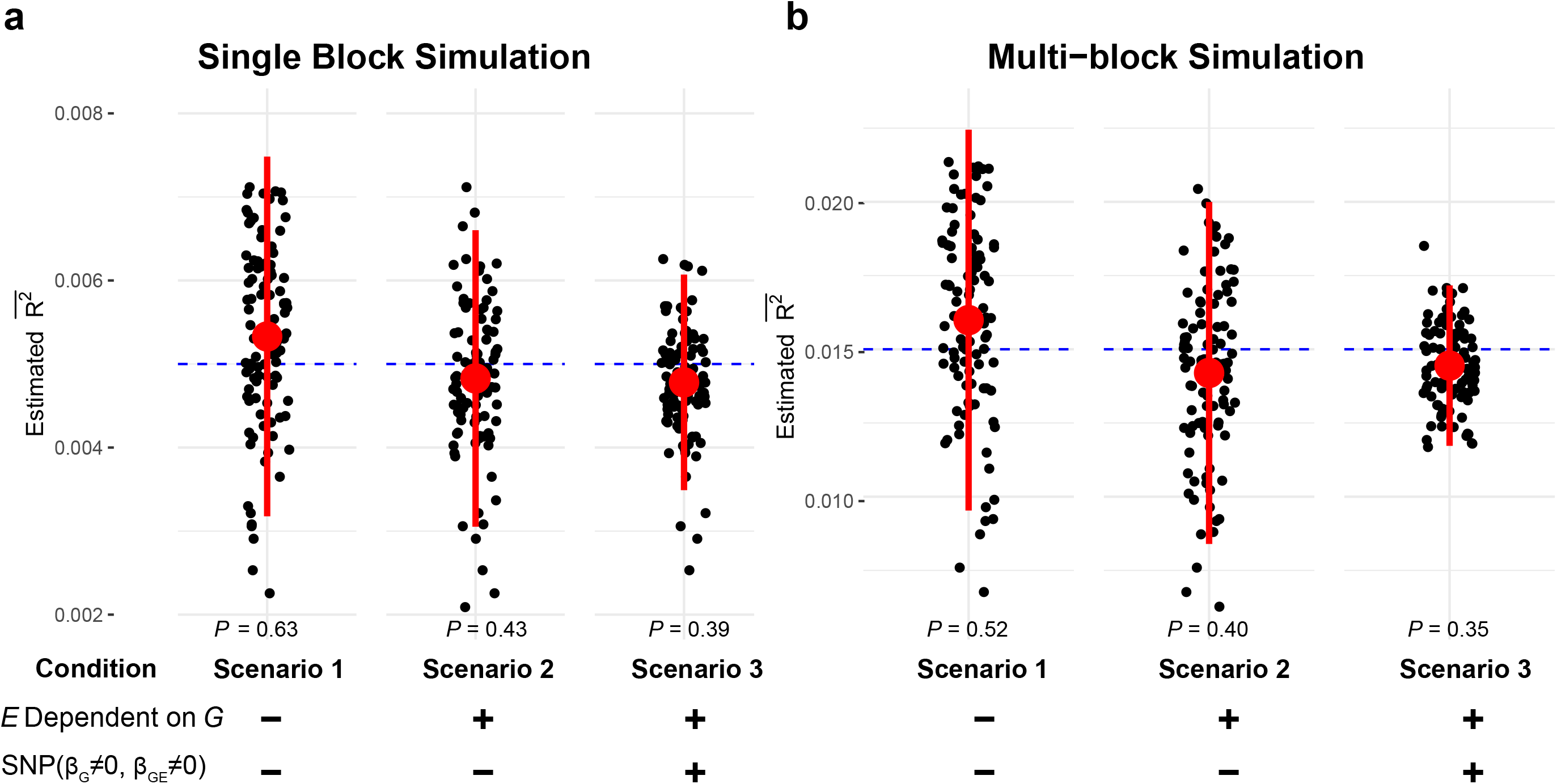
Estimation of variance explained by GxE interactions for 100 simulated phenotypes. Estimation of variance explained by GxE interactions under three simulation scenarios. (+) indicates that the presence of a specific condition, while (-) indicates the absence of a condition. The “*E* dependent on *G”* condition denotes the case where environment effect SNPs are a subset of the same genetic effect SNPs. The “SNP ((β_*G,i*_ ≠ 0, β_*GE,i*_ ≠ 0))” condition denotes the case where a single SNP has both non-zero genetic and interaction effects. Dashed blue lines denote the true variance set by simulations. **a**, Estimation of variance explained by GxE interactions using a single block in chromosome 18 in three scenarios. **b**, Estimation of variance explained by GxE interactions under the three multi-block simulation scenarios for chromosome 11 (3 blocks). 95% CIs were calculated for simulations as described in the methods. *P*-values were derived via *Z*-test.

### Estimation of Genome-Wide Environmental Interaction Effects

Next, we applied MonsterLM to estimate the variance explained by interactions between waist-hip-ratio (WHR) and genetic variants for eight blood biomarkers (Apolipoprotein B, Bilirubin, Total Cholesterol, CRP, HbA1c, HDL-Cholesterol, LDL-Cholesterol, Triglycerides) linked to cardio-metabolic diseases. WHR was selected as the environmental exposure because it is a measure of central obesity linked to a wide range of adverse metabolic consequences, including diabetes and cardiovascular disease (CVD) ^25^. As such, it represents an excellent marker of the effect of the modern obesogenic environment on metabolism. We observed significant variance explained by interaction effects for five of the eight biomarkers, with interaction 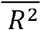 ranging from 0.11 to 0.58 (Figure 3A). As expected, all heritability estimates were significant and consistent with previous work ^16^. Furthermore, we observed the presence of significant directionality for interaction effects at both *P*_*G*_ and *P*_*GE*_ <10^−3^ significance threshold (Figure 3B; Supplementary Figure 1). When stratifying variants according to MAF and LDscore, there was a general tendency for SNPs with low MAF (i.e. 0.01 < MAF < 0.1) and higher LDscore to disproportionally contribute to interaction variance explained per SNP (Supplementary Figure 2)

**Figure 3.**
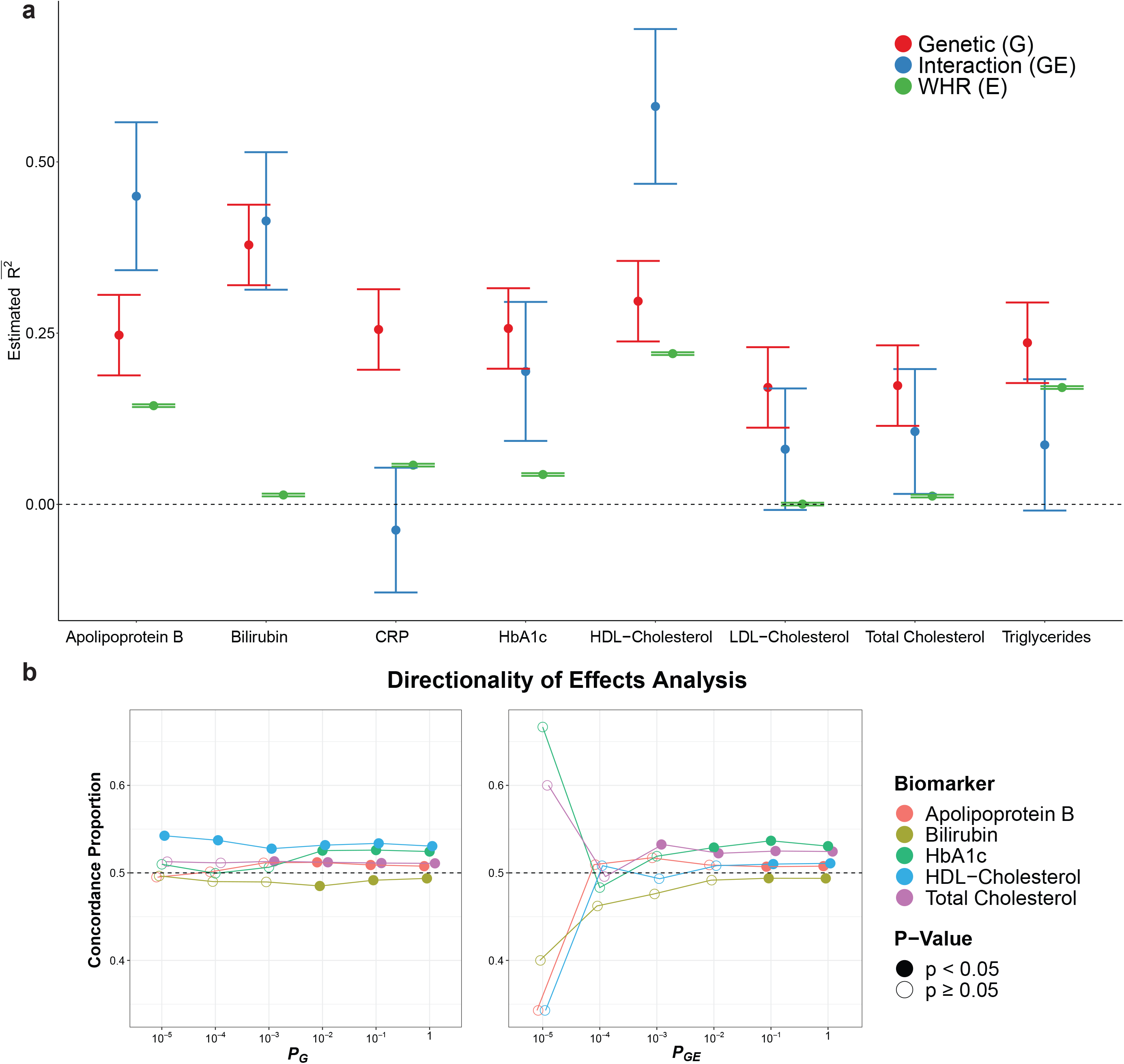
**Estimates of genetic, interaction, and environment (WHR) 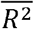 for eight biomarkers and associated directionality of effects.** Studied biomarkers were residualized for age, sex, WHR and the first 20 genetic principal components. Phenotypes were quantile normalized and mean imputed as per methods. 95% CIs were calculated for each estimate as described in the online methods. **a**, Genetic, interaction, and environment (WHR) variance estimated 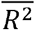 for each biomarker using the MonsterLM protocol. **b**, The directionality of effects for derived interaction estimates. SNPs were filtered based on univariate *P*_*G*_, *P*_*GE*_ and LD (r^2^ < 0.1) for each biomarker. Directionality is concordant when 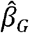 and 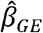 have the same sign (+/+, -/-) and discordant when they have opposite signs (+/-, -/+). Two-proportion *Z*-tests were used to compare each directionality result with a null value of 0.5.

The presence of significant gene-by-WHR (GxWHR) interactions prompted additional questions. First, do GxE interactions arise from SNPs strongly associated with the trait of interest, as has been commonly assumed, or are the variants contributing to GxE interactions independent from those with marginal effects? To address this question, we randomly split participants into a discovery set comprising 80% of participants (260,792 individuals) with the remaining 20% comprising the validation set. Using the five biomarkers with significant GxE interaction variance, we conducted linear regression on the discovery set using biomarker concentration as the outcome variable and a single SNP as the predictor variable, repeating this process for all SNPs and extracting *P*_*G*_. We then selected SNPs according to six association *P*_*G*_ thresholds: <1 (i.e. all SNPs), < 10^−1^, <10^−2^, <10^−3^, <10^−4^, <10^−5^. Each SNP set was then tested for association with the corresponding biomarker in the validation set, using the least number of blocks possible. We evaluated the total 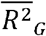 and 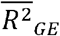 for each of the five SNP sets. The 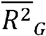 and 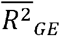 was then compared to the variance explained when including all SNPs (i.e. *P*_*G*_ < 1) for the validation set (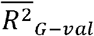 and 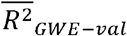). We estimated the proportion of 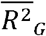 recovered when including an increasing proportion of SNPs in the analysis (Figure 4; Supplementary Figure 3). We observed that between 51-86% of the original 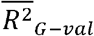 calculated in the validation set could be recovered only using SNPs with *P*_*G*_ <10^−3^ from the discovery set (Supplementary Table 2). We then similarly estimated the proportion of variance explained by GxE interactions recovered when including an increasing proportion of SNPs, based on *P*_*G*_. At a *P*_*G*_ threshold of <10^−3^, only 1-8% of total 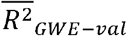 was recovered in the validation set (Figure 4), suggesting that a majority of 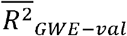 involves SNPs with *P*_*G*_ >10^−3^. At the *P*_*G*_ <10^−2^ threshold, the interaction variance recovered ranged from 2-13% whereas the corresponding range was 0-58% at the *P*_*G*_ <10^−1^ threshold.

**Figure 4.**
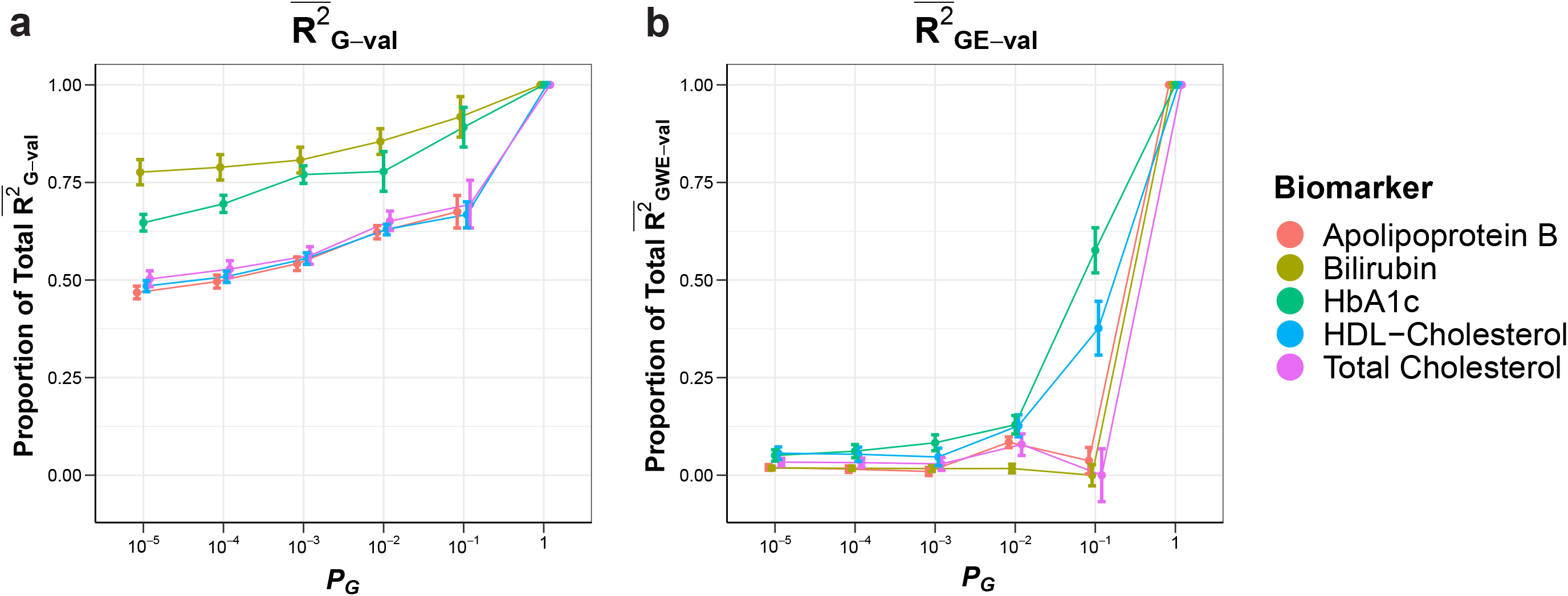
Proportion of 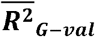 and 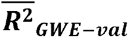 as a function of *P*_*G*_. **a**, The proportion of total 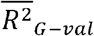 recovered in the validation set at each discovery sample *P*_*G*_ for the five biomarkers with significant interaction variance. **b**, The proportion of total interaction 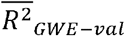 recovered in the validation set at each discovery sample *P*_*G*_ threshold for the same biomarkers. 95% CI were derived based on the upper and lower bounds of each estimate in proportion to either total 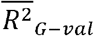 or 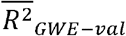.

As our results showed that GxE interactions are largely derived from SNPs without strong marginal associations, we next sought to address whether a few strong GxE interactions are responsible for the large variance explained by interactions, or whether it is the result of many small interactions. We conducted regression on each SNP and its associated interaction from the discovery set. We selected interactions based on five discovery *P*_*GE*_ thresholds: <1 (i.e. all SNPs), <10^−1^, <10^−2^, <10^−3^, <10^−4^, <10^−5^. In other words, an interaction term was included in the validation sample analysis if it passed the *P*_*GE*_ threshold in the discovery set. Importantly, all SNPs were included in the analysis, irrespective of whether their corresponding interaction terms were included or not. The interaction 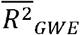 were computed in the validation set and compared to the 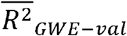 estimates (Figure 5; Supplementary Figure 4). We observed that up to 45% of the total 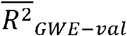 was recovered at a discovery *P*_*GE*_ threshold <10^−3^, corresponding to 0.2-3.3% of the SNPs tested in our initial analyses (Supplementary Figure 4). Indeed, high recovery of variance explained by interaction was also observed at the *P*_*GE*_ <10^−2^ (range: 14-78%) and *P*_*GE*_ <10^−1^ (range: 48-94%) thresholds. To confirm the specificity of interaction effects, we conducted a sensitivity analysis using Apolipoprotein B (Supplementary Table 3). We randomly selected a set of interaction terms equal to the number of interactions included at the *P*_*GE*_ <10^−2^ threshold (62,904 SNPs out of 1.2 million SNPs tested). We then calculated the 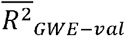 using this set of randomly chosen interaction effects. The randomly selected SNPs had an 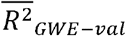 of 0.02, compared to an 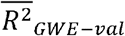 of 0.25 for the interaction terms with *P*_*GE*_ <10^−2^ in the validation set.

**Figure 5.**
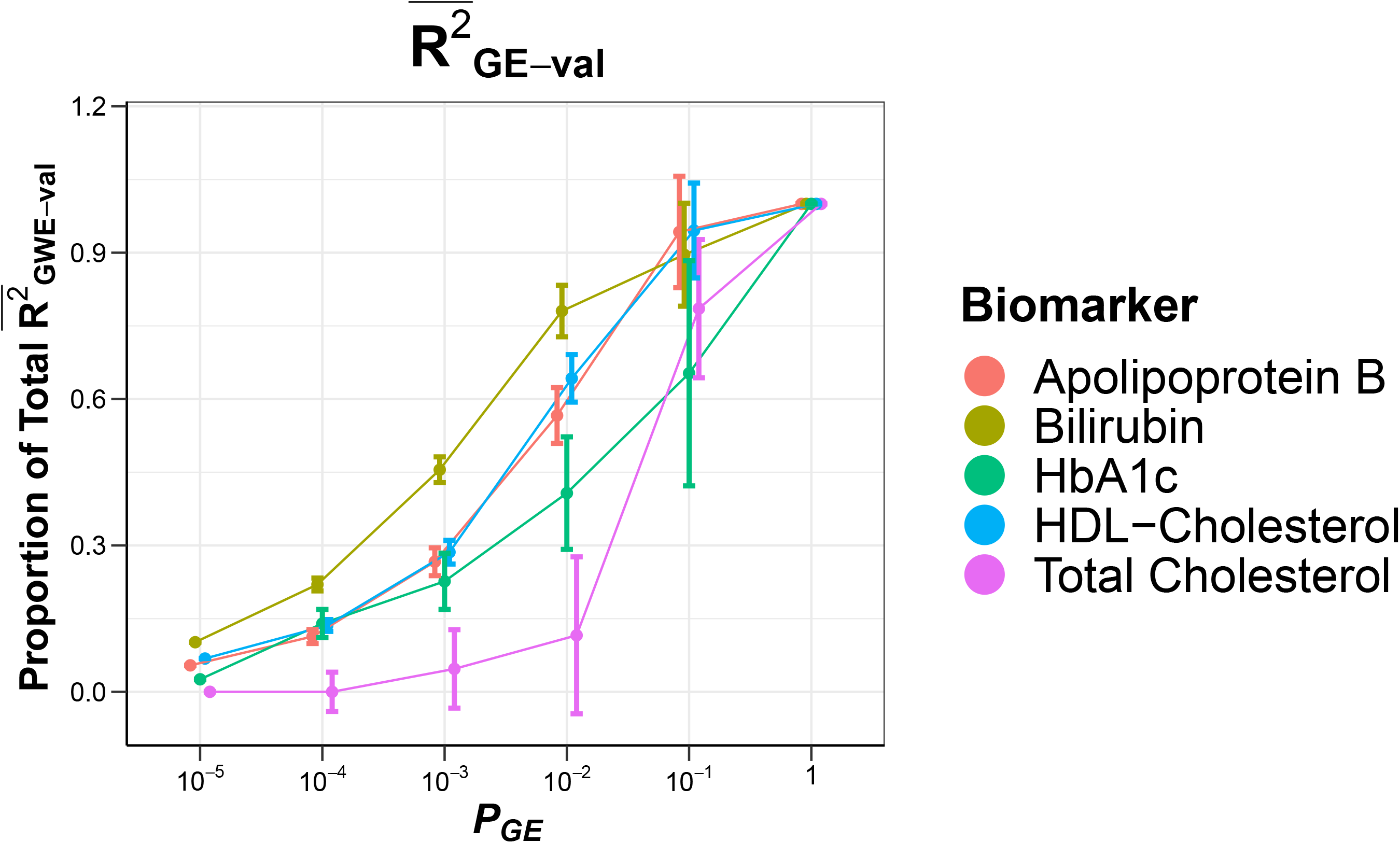
Proportion of 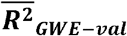 estimates as a function of *P*_*GE*_ thresholds. Proportion of total 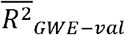 recovered in the validation set at each univariate *P*_*GE*_ threshold for the five biomarkers with significant interaction variance. 95% CI were derived based on the upper and lower bounds of each estimate in proportion to total 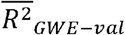.

### Polygenic Scores Analysis

Finally, we examined if the predictiveness of polygenic score (*PS*) could be improved by incorporating interactions. To select SNPs and interaction effects to be included in each *PS*, we used both *P*_*G*_ and *P*_*GE*_ thresholds of 10^−2^, 10^−3^, 10^−4^, and 10^−5^ in the discovery set when testing either each SNP individually or both a single SNP and corresponding interaction, respectively. Each *PS* was then tested in the validation sample for association with its corresponding biomarker. *PS* prediction 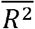 was modestly improved for the four biomarkers with the highest interaction variance by incorporating interaction effects (Figure 6), with the relative increase in Prediction 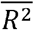 ranging from 0% to 8% across the biomarkers analyzed. Significant improvements in prediction of Apolipoprotein B, Bilirubin and HDL-Cholesterol levels were observed at the 95% confidence level (for interaction significance thresholds of 10^−3^, 10^−4^,10^−5^; Supplementary Table 4). Notably, there was no improvement in the Total Cholesterol *PS* with Interactions (*PS*_*GE*_) compared to their respective *PS* without interactions (*PS*_*G*_) values (Figure 6), consistent with the *P*_*GE*_ results for Total Cholesterol (Figure 5).

**Figure 6.**
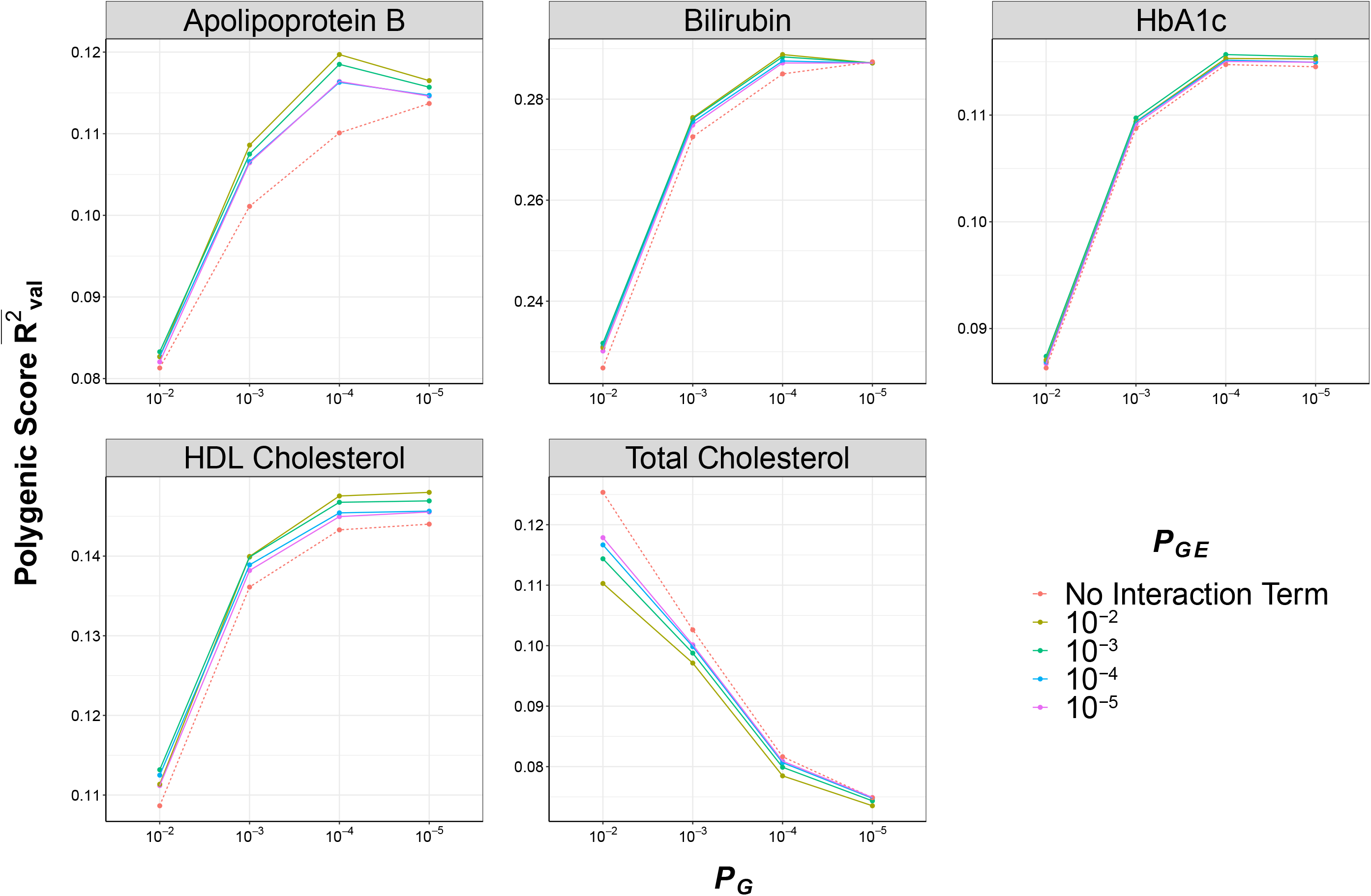
Polygenic score prediction 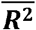 with and without incorporation of interaction effects. For each biomarker, there are 20 different conditions based on discovery sample *P*_*G*_ and *P*_*GE*_ thresholds. The polygenic score 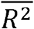 was estimated in the validation sample based on discovery sample 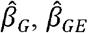 values.

## Discussion

In this report, we developed a novel method, MonsterLM, to estimate variance explained by genome-wide interactions with environmental exposures. Using simulations, we verified that MonsterLM estimates the variance explained by interaction effects accurately and precisely. Analysis of UK Biobank biomarker data demonstrated the presence of significant GxE interactions effects with WHR, a marker of metabolically deleterious adiposity. The interaction estimates for five of the eight biomarkers analysed were significant with estimates ranging between 0.11 to 0.58 of overall variance, prompting further analyses into these results.

MonsterLM provides distinct advantages over current methods for GxE analysis (Table 1) ^26–30^. In most settings, inference methods for genome-wide SNP-heritability and GxE interactions make assumptions on genetic architecture. These assumptions are parametrized by polygenicity (the number of variants with effects) and MAF/LD-dependence (the coupling of effects with MAF, LD or other functional annotations). Since the true genetic architecture of any given trait is unknown, existing methods are susceptible to bias and often yield vastly different estimates even when applied to the same data^10–12^. This is also the case for the estimation of Genome-Wide Environment interactions, where different assumptions about the structure of interactions result in a variety of different estimates^26–30^. Although multi-component methods that stratify SNPs by LD/MAF can address these robustness issues, fitting multiple variance components to biobank scale data is highly resource intensive^16^, and this problem is compounded when considering interactions where the number of variables analyzed increases by two-fold. Alternate methods that explicitly model these dependencies are also sensitive to model misspecification ^9–13^. MonsterLM makes no assumption with respect to the genetic model and does not rely on parametrization for underlying assumptions. Our partitioning approach combined with methods to accelerate computations allows for fast, unbiased genome-wide computations of heritability and GxE interactions for both small datasets and biobank-scale data. Our method also enables testing for interactions with specific environmental exposures instead of overall effects from multiple environmental outcomes. By reducing the assumptions required for computing heritability and GxE interactions, MonsterLM has the potential to uncover greater insights into the genetic architecture of GxE interactions.

Our analyses revealed the presence of significant GxE interactions for five of eight blood biomarkers with WHR. Interaction effects ranged from null to very strong, and in the cases of Apolipoprotein B, Bilirubin, and HDL-Cholesterol, explained a higher proportion of overall variance than heritability. These results have important implications for future research. First, our observations suggest that there are real interactions between genetics and exposures that contribute greatly to complex trait variance. Second, genetic associations are likely to be heterogenous when comparing populations with dramatically different obesogenic environmental exposures. The observation that GxE effects do not come from SNPs with strong marginal effects suggests this may not impact top GWAS hits excessively. We also observed the presence of significant directionality effects for strongly significant SNPs and their associated interaction effects, which suggest an overall greater impact of genetic variation under certain environmental conditions. There are also clinical implications for these observations. For instance, Apolipoprotein B is a *bona fide* risk factor for coronary artery disease (CAD) ^31^. A strong interaction effect with WHR is observed, suggesting WHR is also an important modulator of genetic risk of CAD mediated through Apolipoprotein B.

Our results also provide some insights into why identification of GxE interactions has been challenging^1^. Many prior studies have reasonably focused the search for significant GxE interactions on variants with genome-wide significant marginal effects. However, our results show that only a small proportion of GxE interaction effects can be explained by such variants. Rather, the majority of GxE interaction effects are due to variants with unremarkable marginal effects. On the other hand, we also show that a relatively small minority of variants is responsible for a disproportionate contribution to GxE interactions. Altogether, these findings offer hope that the identification of specific interactions is possible. Indeed, we also show in a proof-of-concept experiment that incorporation of GxE interactions can significantly improve PS prediction, albeit modestly.

Some limitations are worth mentioning. First, we quantile normalized all traits before analysis, and while this protects against potential scaling effects, it could also bias results towards the null. Second, MonsterLM is not meant to identify specific GxE interactions but rather to quantify the overall, genome-wide contributions of GxE interactions to continuous traits. Another limitation includes the potential loss of information from LD pruning to account for high correlation in the genotype data and from filtering rare variants (MAF<1%).

In this report, we have established the presence of GxE interactions in cardiometabolic biomarkers. We observed that SNPs with strong marginal effects contribute weakly to the variance of GxE interaction effects, and that there is a disproportionate contribution from a relatively small minority of variants. Our results also highlight the potential for pathway analysis, examining specific genes involved in GxE interactions. MonsterLM provides flexibility for any form of genetic architecture, environmental exposures and interaction models, and serves as the basis for more advanced future analyses into the specifics of genome-wide environmental interactions and importantly, the contribution of GxE interactions to dichotomous traits such as disease status.

## Data Availability

All code is available upon request.

## Acknowledgements

The authors are thankful for all the UK Biobank participants.

## Author Contributions

**MK**: data curation, software, formal analysis, investigation, visualization, writing (original draft); **MD**: formal analysis, visualization, writing (review and editing); **CJ** : formal analysis, visualization, writing (review and editing); **NP**: formal analysis; **MC**: data curation, analysis interpretation, writing (review and editing); **SM**: data curation, software; **SD**: software; **WN**: software **JP**: software; **GP**: conceptualization, supervision, funding acquisition, methodology, project administration, writing (review and edit).

## Code Availability Statement

All custom code is available upon request.

## References

1. Aschard, H. A perspective on interaction effects in genetic association studies. Genet. Epidemiol. 40, 678–688 (2016).

2. Dempfle, A. et al.. Gene-environment interactions for complex traits: definitions, methodological requirements and challenges. Eur. J. Hum. Genet. EJHG 16, 1164–1172 (2008).

3. Castaldi, P. J. et al.. Screening for interaction effects in gene expression data. PloS One 12, e0173847 (2017).

4. Kim, J. et al.. Joint Analysis of Multiple Interaction Parameters in Genetic Association Studies. Genetics 211, 483–494 (2019).

5. Dai, J. Y. et al.. Simultaneously testing for marginal genetic association and gene-environment interaction. Am. J. Epidemiol. 176, 164–173 (2012).

6. Patel, C. J., Chen, R., Kodama, K., Ioannidis, J. P. A. & Butte, A. J. Systematic identification of interaction effects between genome- and environment-wide associations in type 2 diabetes mellitus. Hum. Genet. 132, 495–508 (2013).

7. Almasy, L. & Blangero, J. Variance component methods for analysis of complex phenotypes. Cold Spring Harb. Protoc. 2010, pdb.top77 (2010).

8. Veerman, J. R., Leday, G. G. R. & Wiel, M. A. van de. Estimation of variance components, heritability and the ridge penalty in high-dimensional generalized linear models. Commun. Stat. - Simul. Comput. 0, 1–19 (2019).

9. Speed, D., Hemani, G., Johnson, M. R. & Balding, D. J. Improved Heritability Estimation from Genome-wide SNPs. Am. J. Hum. Genet. 91, 1011–1021 (2012).

10. Speed, D. et al.. Reevaluation of SNP heritability in complex human traits. Nat. Genet. 49, 986–992 (2017).

11. Speed, D. & Balding, D. J. SumHer better estimates the SNP heritability of complex traits from summary statistics. Nat. Genet. 51, 277–284 (2019).

12. Evans, L. M. et al.. Comparison of methods that use whole genome data to estimate the heritability and genetic architecture of complex traits. Nat. Genet. 50, 737–745 (2018).

13. Gazal, S., Marquez-Luna, C., Finucane, H. K. & Price, A. L. Reconciling S-LDSC and LDAK models and functional enrichment estimates. http://biorxiv.org/lookup/doi/10.1101/256412 (2018) doi:10.1101/256412.

14. Yang, J. et al.. Genetic variance estimation with imputed variants finds negligible missing heritability for human height and body mass index. Nat. Genet. 47, 1114–1120 (2015).

15. Bycroft, C. et al.. The UK Biobank resource with deep phenotyping and genomic data. Nature 562, 203–209 (2018).

16. Schizophrenia Working Group of the Psychiatric Genomics Consortium et al. LD Score regression distinguishes confounding from polygenicity in genome-wide association studies. Nat. Genet. 47, 291–295 (2015).

17. Finucane, H. K. et al.. Partitioning heritability by functional annotation using genome-wide association summary statistics. Nat. Genet. 47, 1228–1235 (2015).

18. Gazal, S. et al.. Linkage disequilibrium-dependent architecture of human complex traits shows action of negative selection. Nat. Genet. 49, 1421–1427 (2017).

19. Hou, K. et al.. Accurate estimation of SNP-heritability from biobank-scale data irrespective of genetic architecture. Nat. Genet. 51, 1244–1251 (2019).

20. de Los Campos, G., Hickey, J. M., Pong-Wong, R., Daetwyler, H. D. & Calus, M. P. L. Whole-genome regression and prediction methods applied to plant and animal breeding. Genetics 193, 327–345 (2013).

21. Mayhew, A. J. & Meyre, D. Assessing the Heritability of Complex Traits in Humans: Methodological Challenges and Opportunities. Curr. Genomics 18, 332–340 (2017).

22. Browning, S. R. & Browning, B. L. Population structure can inflate SNP-based heritability estimates. Am. J. Hum. Genet. 89, 191–193; author reply 193-195 (2011).

23. Shewchuk, J. R. An Introduction to the Conjugate Gradient Method Without the Agonizing Pain. (1994).

24. Nogueira, B. & Pinheiro, R. G. S. A GPU based local search algorithm for the unweighted and weighted maximum s-plex problems. Ann. Oper. Res. 284, 367–400 (2020).

25. Poppitt, S. D. et al.. Long-term effects of ad libitum low-fat, high-carbohydrate diets on body weight and serum lipids in overweight subjects with metabolic syndrome. Am. J. Clin. Nutr. 75, 11–20 (2002).

26. Moore, R. et al.. A linear mixed model approach to study multivariate gene-environment interactions. Nat. Genet. 51, 180–186 (2019).

27. Robinson, M. R. et al.. Genotype-covariate interaction effects and the heritability of adult body mass index. Nat. Genet. 49, 1174–1181 (2017).

28. Dahl, A. et al.. A Robust Method Uncovers Significant Context-Specific Heritability in Diverse Complex Traits. Am. J. Hum. Genet. 106, 71–91 (2020).

29. Sulc, J. et al.. Quantification of the overall contribution of gene-environment interaction for obesity-related traits. Nat. Commun. 11, (2020).

30. Kerin, M. & Marchini, J. Inferring Gene-by-Environment Interactions with a Bayesian Whole-Genome Regression Model. Am. J. Hum. Genet. 107, 698–713 (2020).

31. Sniderman, A. D. et al.. Apolipoprotein B Particles and Cardiovascular Disease: A Narrative Review. JAMA Cardiol. 4, 1287–1295 (2019).

32. Sudlow, C. et al.. UK Biobank: An Open Access Resource for Identifying the Causes of a Wide Range of Complex Diseases of Middle and Old Age. PLOS Med. 12, e1001779 (2015).

33. De La Vega, F. M. & Bustamante, C. D. Polygenic risk scores: a biased predictionã Genome Med. 10, 100 (2018).

34. Graf, R. G. & Alf, E. F. Correlations Redux: Asymptotic Confidence Limits for Partial and Squared Multiple Correlations. Appl. Psychol. Meas. 23, 116–119 (1999).

