## Supplementary material for "A versatile, fast and unbiased method for estimation of gene-by-environment interaction effects on biobank-scale datasets": GxE interaction effects Supplementary Appendix.docx

### Supplementary Materials

#### Supplementary Table 1 – Comparing the CG method with GPU acceleration to the Least Squares Inversion for Beta

Attached as supplementary materials (csv).

Supplementary Table 2 – Validation Set Estimates

Attached as supplementary materials (csv).

#### Supplementary Table 3 – Sensitivity Analysis Table

Attached as supplementary materials (csv).

#### Supplementary Table 4 – PS Confidence Intervals

Attached as supplementary materials (csv).

### Supplementary Figure Legends

**Supplementary Figure 1 |** Number of SNPs included in the directionality analysis by $P_{G}$ and $P_{GE}$ thresholds for each biomarker.

**Supplementary Figure 2 |** Estimated $\overline{R^{2}}$ per SNP according to MAF and LD categories for the five biomarkers with significant interaction variance.

**Supplementary Figure 3 |** Number of SNPs included by $P_{G}$ threshold for each biomarker in the proportion of genetic variance analysis.

**Supplementary Figure 4 |** Number of SNPs included by $P_{GE}$ threshold for each biomarker in the proportion of interaction variance analysis.
