## Supplementary figures and images for "A versatile, fast and unbiased method for estimation of gene-by-environment interaction effects on biobank-scale datasets"

### Supp_Fig1.pdf

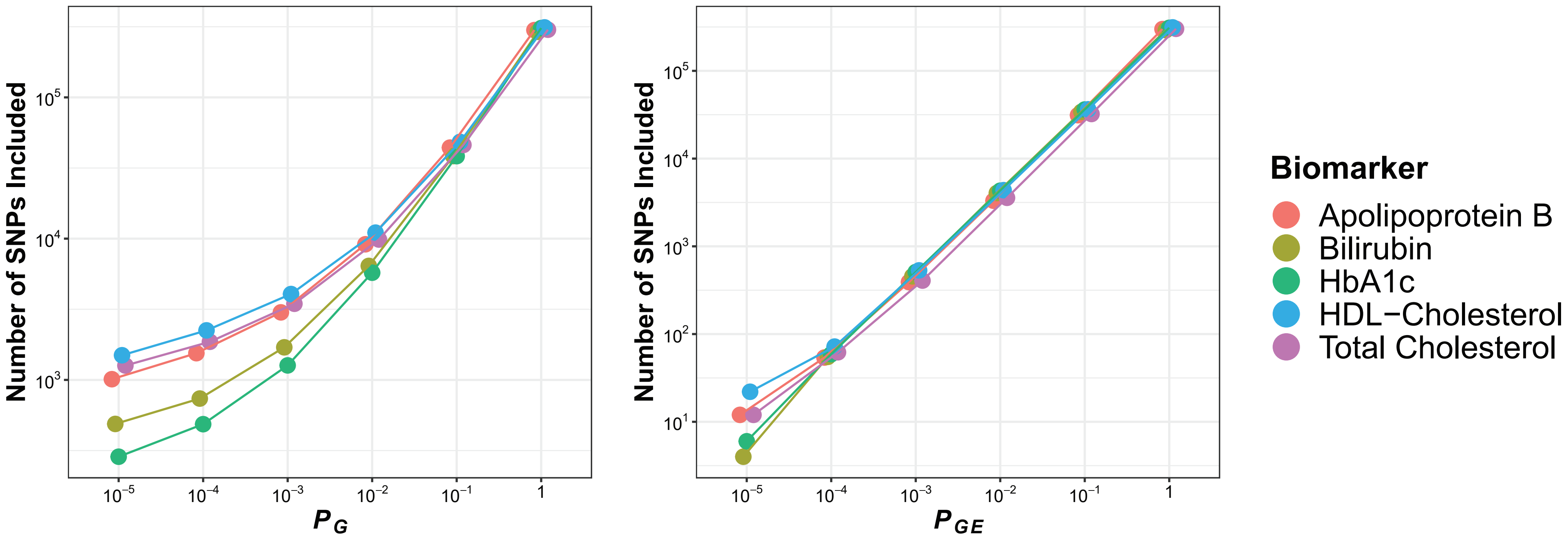

### Supp_Fig2.pdf

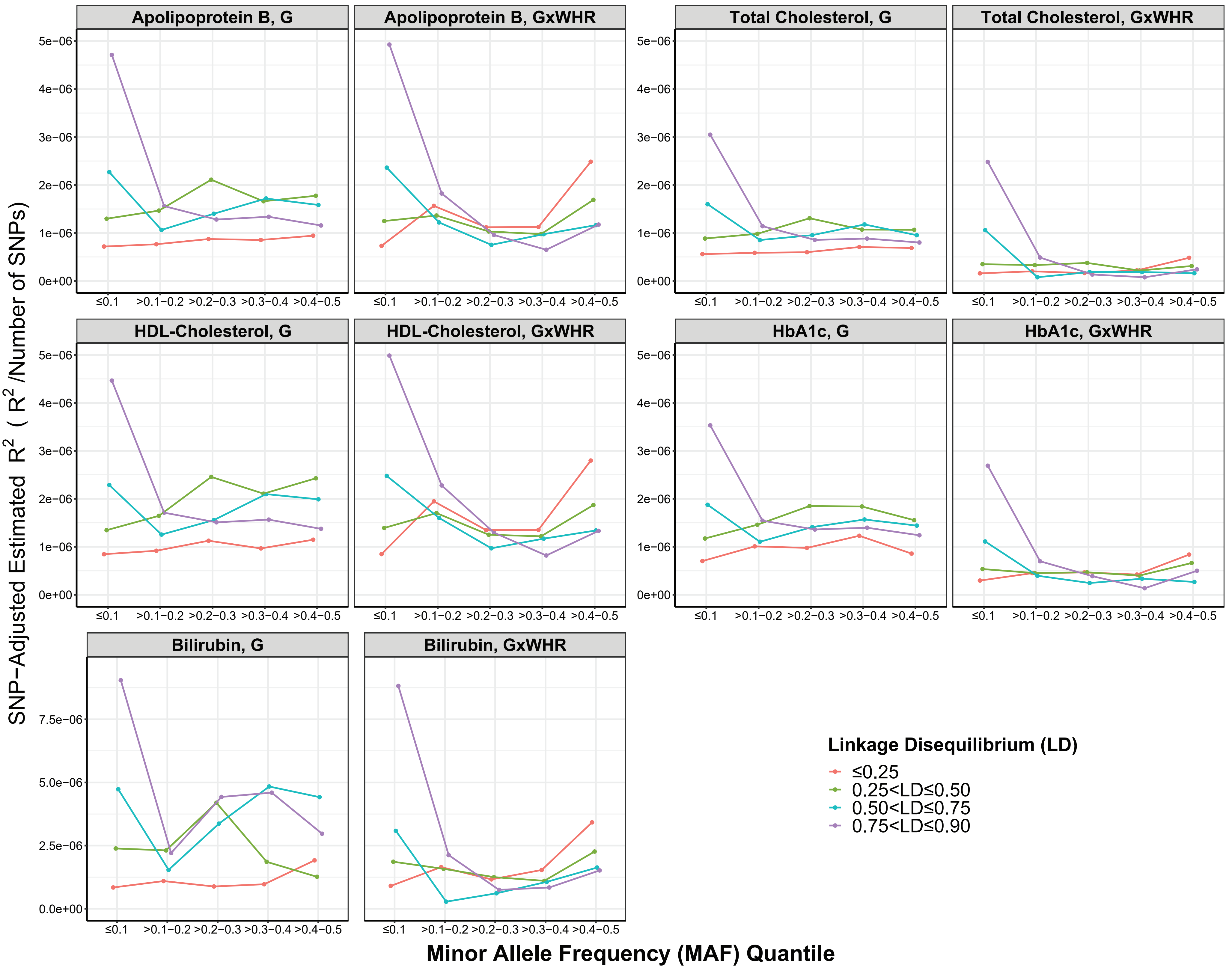

### Supp_Fig3.pdf

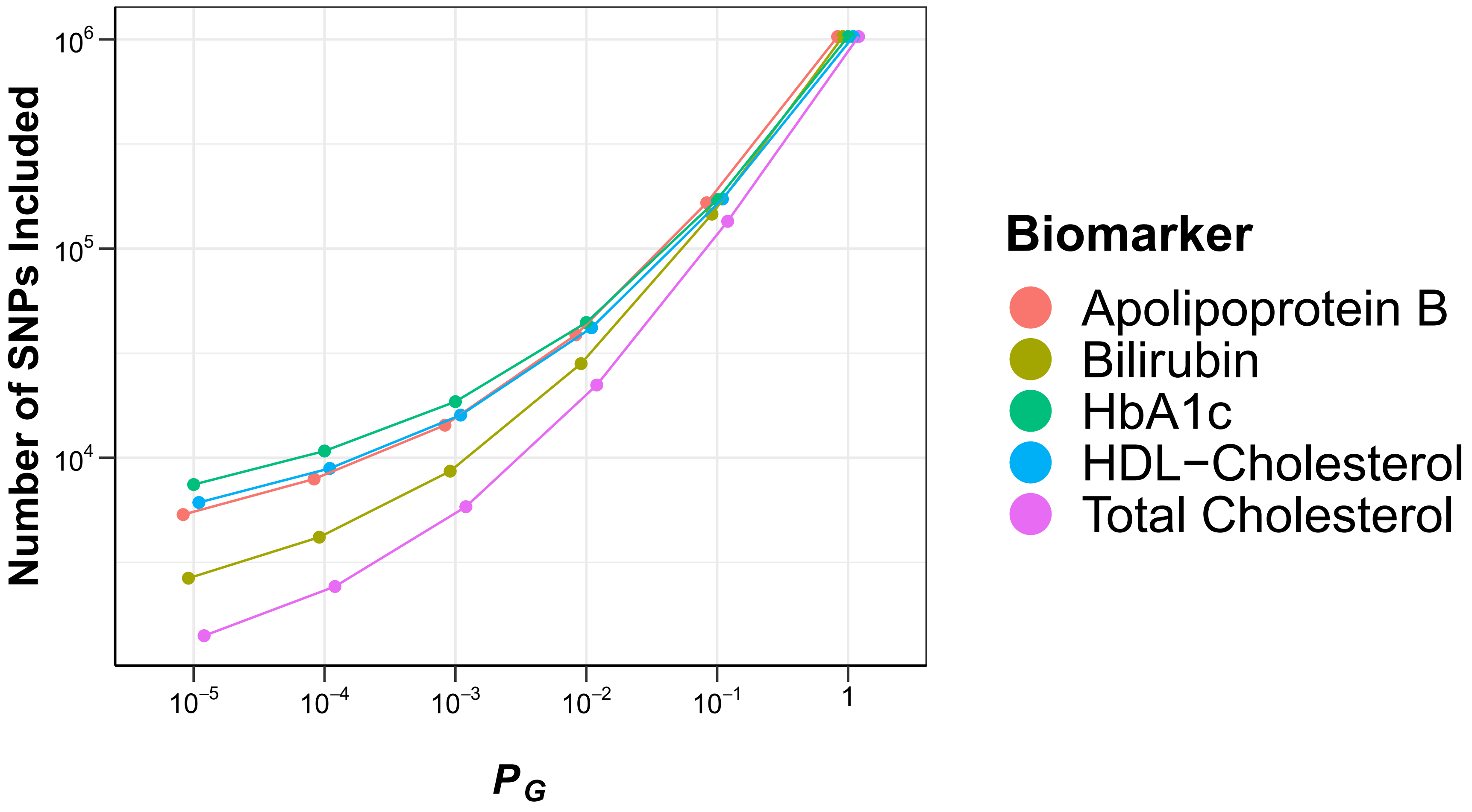

### Supp_Fig4.pdf

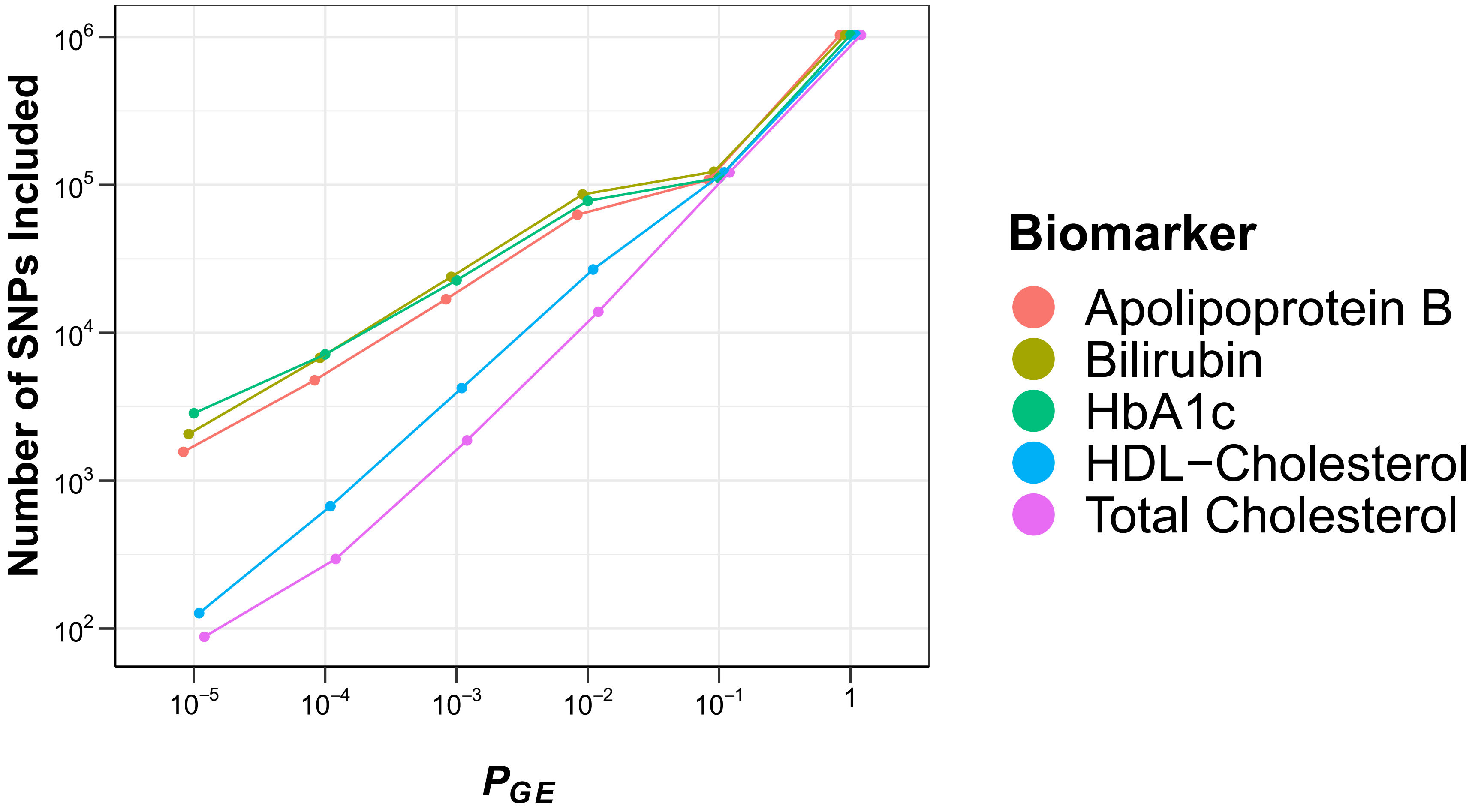

### Supp_Figures_merged.pdf

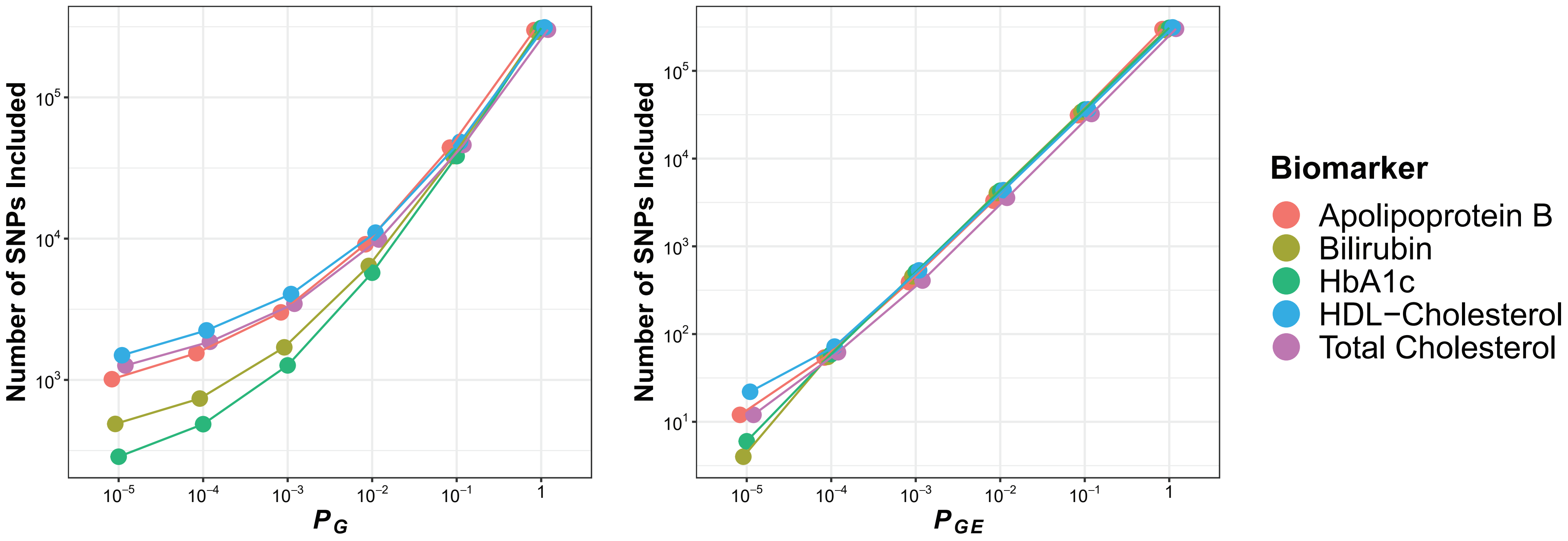

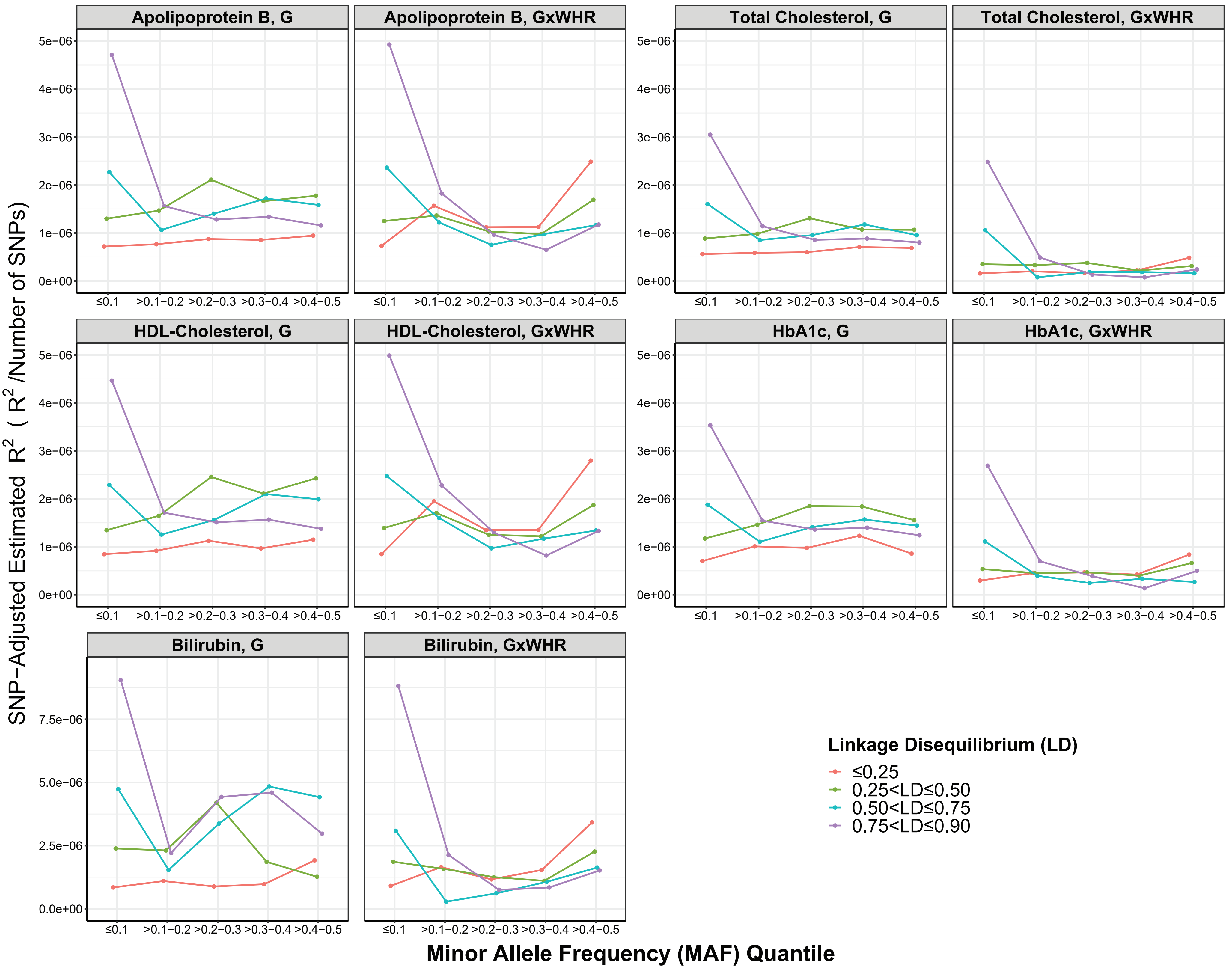

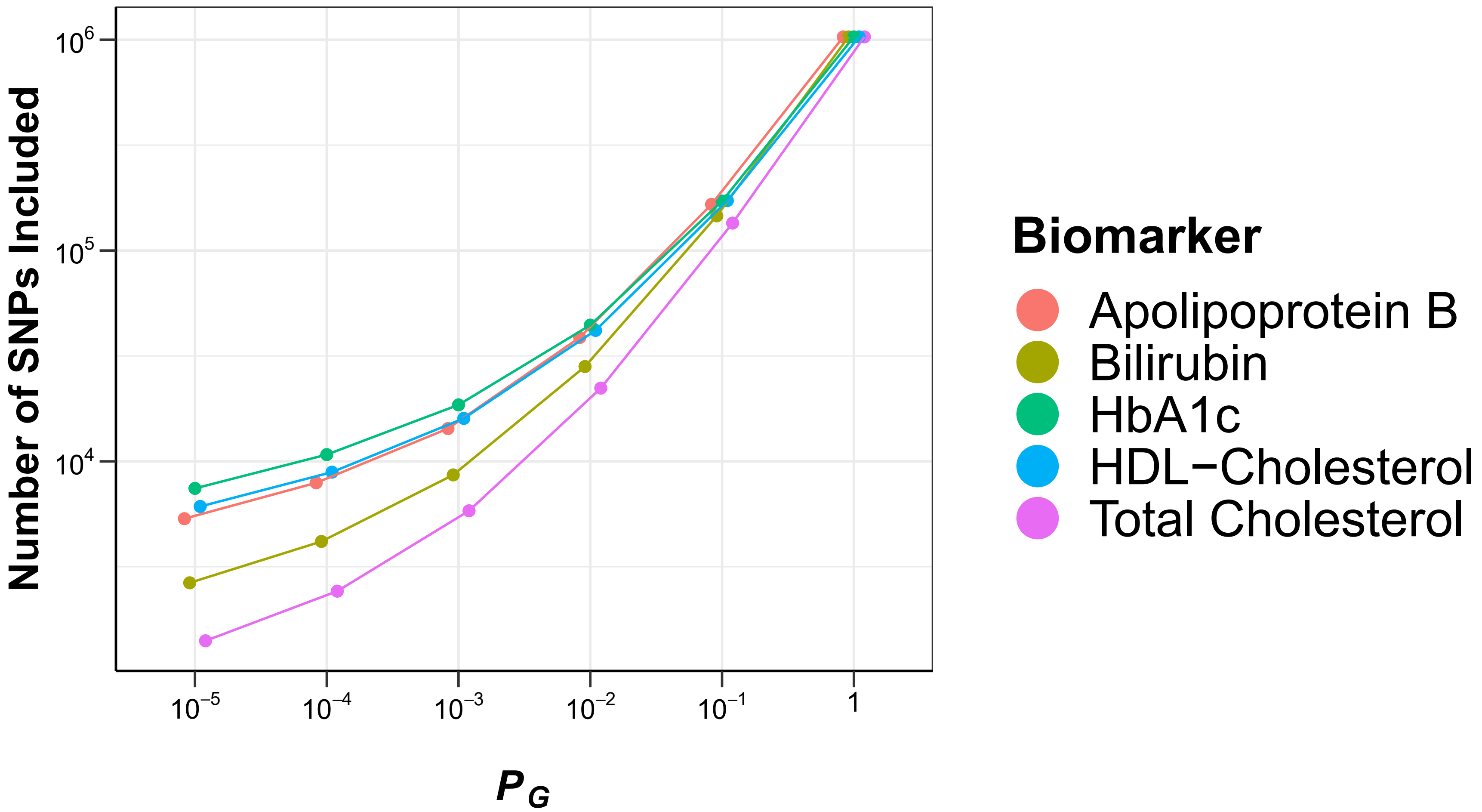

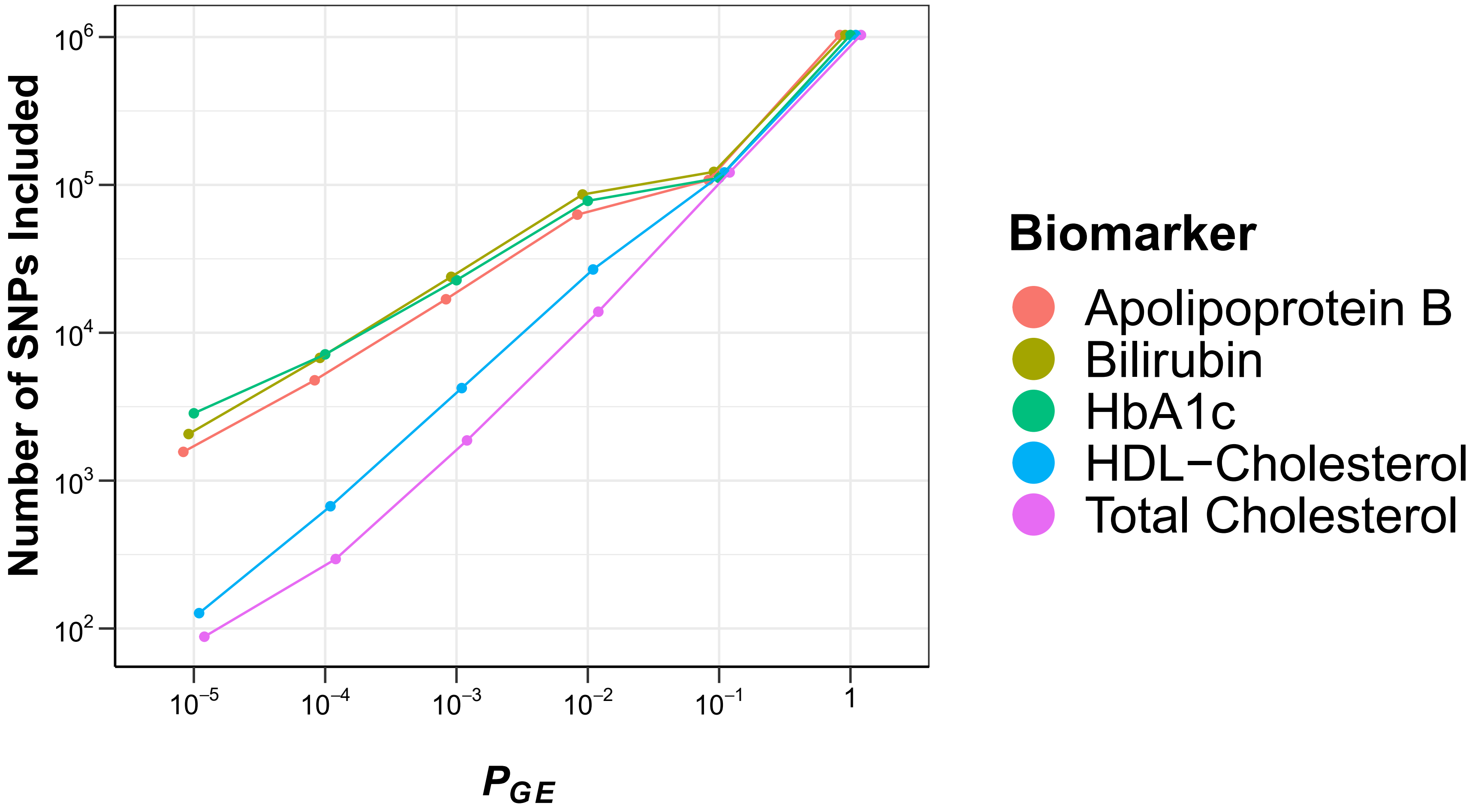
